# *ACTR1A* has pleiotropic effects on risk of leprosy, inflammatory bowel disease and atopy

**DOI:** 10.1101/2022.01.31.22270046

**Authors:** James J Gilchrist, Kathryn Auckland, Tom Parks, Alexander J Mentzer, Lily Goldblatt, Vivek Naranbhai, Gavin Band, Kirk A Rockett, Ousmane B Toure, Salimata Konate, Sibiri Sissoko, Abdoulaye A Djimdé, Mahamadou A Thera, Ogobara K Doumbo, Samba Sow, Sian Floyd, Jörg M Pönnighaus, David K Warndorff, Amelia C Crampin, Paul EM Fine, Benjamin P Fairfax, Adrian VS Hill

## Abstract

Leprosy is a chronic infection of the skin and peripheral nerves caused by *Mycobacterium leprae*. Despite recent improvements in disease control, leprosy remains an important cause of infectious disability globally. Large-scale genetic association studies in Chinese, Vietnamese and Indian populations have identified over 30 susceptibility loci for leprosy. There is a significant burden of leprosy in Africa, however it is uncertain whether the findings of published genetic association studies are generalizable to African populations. To address this, we conducted a genome-wide association study (GWAS) of leprosy in Malawian (327 cases, 436 controls) and Malian (247 cases, 368 controls) individuals. In that analysis, we replicated five risk loci previously reported in China, Vietnam and India; MHC Class I and II, *LACC1* (2 independent loci) and *SLC29A3*. We further identified a novel leprosy susceptibility locus at 10q24 (rs2015583: combined *p* = 8.81 × 10^−9^; *OR* = 0.51 [95% CI 0.40 *-* 0.64]). The leprosy risk locus is a determinant of *ACTR1A* RNA expression in CD4^+^ T cells (posterior probability of colocalization - *PP* = 0.96). Furthermore, it demonstrates pleiotropy with established risk loci for inflammatory bowel disease and atopic disease. Reduced *ACTR1A* expression decreases susceptibility to leprosy and atopy but increases risk of inflammatory bowel disease. A shared genetic architecture for leprosy and inflammatory bowel disease has been previously described. We expand on this, strengthening the evidence that selection pressure driven by leprosy has shaped the evolution of autoimmune and atopic disease in modern populations. More broadly, our data highlights the importance of defining the genetic architecture of disease across genetically diverse populations, and that disease insights derived from GWAS in one population may not translate to all affected populations.

**Author Summary:** Leprosy remains a leading cause of infectious disability globally. Human genetic variation is a major determinant of susceptibility to infection, including leprosy. Large-scale genetic association studies have been pivotal in advancing our understanding of leprosy biology. These studies have been performed in Chinese, Vietnamese and Indian populations, and it remains unclear whether these insights are informative of leprosy susceptibility in African populations. To address this, we performed a genome-wide association study of leprosy susceptibility in Malawi and Mali. In doing so we replicate known leprosy susceptibility loci at MHC class I and II, *LACC1* and *SLC29A3*. Furthermore, we identify a novel leprosy susceptibility locus, which modifies expression of *ACTR1A* in CD4^+^ T cells and demonstrates pleiotropy with inflammatory bowel disease (IBD) and atopic disease. These data deepen our understanding of leprosy biology, identifying *ACTR1A* expression in CD4^+^ T cells as a determinant of leprosy disease risk, and further define the role of this ancient pathogen in the evolution of immune-mediated diseases in modern populations.

## Introduction

Leprosy is a chronic infectious disease affecting the skin and peripheral nerves caused by *Mycobacterium leprae*. It is a leading infectious cause of disability (Britton & Lockwood, 2004). The introduction of multidrug therapy (“Chemotherapy of leprosy. Report of a WHO Study Group.”, 1994), widespread use of BCG vaccination (Pönnighaus et al., 1992), and the 1991 World Health Assembly resolution to eliminate leprosy by the year 2000 have all contributed to a decline in disease burden; nevertheless, over 200,000 new cases continue to be reported annually (https://www.who.int/gho/neglected_diseases/leprosy/en/), numbers which are likely to represent a considerable underestimate of the true disease burden (Smith et al., 2015).

Large-scale, unbiased genetic association studies in Chinese (Liu et al., 2015; Liu et al., 2017; Wang et al., 2018; Wang et al., 2016; F.-R. Zhang et al., 2009; F. Zhang et al., 2011), Indian (Wong et al., 2010) and Vietnamese (Gzara et al., 2020) populations have identified and validated 34 genetic loci independently associated with leprosy outside the HLA region, as well as independent HLA class I and class II associations. A key feature of these studies has been the demonstration of considerable genetic heterogeneity in leprosy susceptibility between populations. For instance, while genetic variation at *TLR1* associates with leprosy risk in Indian and Turkish populations, this finding has not been replicated in Chinese and Vietnamese populations. To date, there are no published genome-wide association studies (GWAS) of leprosy in African populations. This is important as there remains a significant burden of leprosy in Africa and the observed inter-population heterogeneity reported in the Chinese, Vietnamese and Indian studies suggest that the published GWAS findings may not be generalizable to African populations. To address this, we have performed a GWAS of leprosy susceptibility in Malawian and Malian individuals.

## Results

### Leprosy genome-wide association study

Individuals with leprosy were recruited to the study following clinical and microbiological evaluation within the Karonga Prevention Study (KPS), Karonga, Malawi. Healthy controls were recruited from the same population. Following quality control and genome-wide imputation, genotypes at 10,511,695 loci from 612 samples (284 cases, 328 controls) were included in the association analysis. Inspection of the QQ plot (Supplementary Fig. 1) and the genomic inflation parameter (*λ* = 1.0333) demonstrates that inclusion of the six major principal components as covariates in the model adequately controls for confounding variation. In that analysis, we identified 142 loci, representing 38 independent genomic loci, with suggestive evidence of association (*p* < 1 × 10^−5^) with leprosy in Malawian populations (Supplementary Fig. 2, Supplementary Table 1).

### GWAS replication and meta-analysis

We sought to replicate evidence for leprosy association observed in Malawi among leprosy cases and healthy controls in Mali. Individuals with leprosy were recruited to the study at Mali’s former national leprology centre, Institut Marchoux, (now Hôpital Dermatologique de Bamako), Bamako. Healthy controls were recruited from the same population. Among the Malian samples, 10,514,676 loci and 519 individuals (208 cases, 311 controls) passed QC filters. The QQ plot (Supplementary Fig. 1) and genomic inflation parameter (*λ* = 1.0498) of genome-wide association analysis in the Malian samples demonstrates control of confounding variation with inclusion of the major six major principal components and genotyping platform as covariates in the model. We combined evidence for leprosy association in Malawi and Mali using a fixed-effects meta-analysis (Fig. 1). Of the 142 leprosy-associated loci identified in the discovery analysis, 18 SNPs, at a single genomic locus at 10q24.32 (Fig. 2A), showed evidence of replication in Mali (*p* < 0.05) and overall evidence of association with leprosy exceeding genome-wide significance (*p* < 5 × 10^−8^). The variant with the strongest evidence for leprosy association at that locus is rs2015583: *p* = 8.81 × 10^−9^, *OR* = 0.51 (95% CI 0.40 *-* 0.64). There is no evidence for heterogeneity of effect between populations at rs2015583 (heterogeneity *p* = 0.871, Fig. 2B), and the data best supports a model in which rs2015583 modifies risk of both paucibacillary and multibacillary leprosy (log10 Bayes factor = 6.01, Fig. 2B).

**Figure 1:**
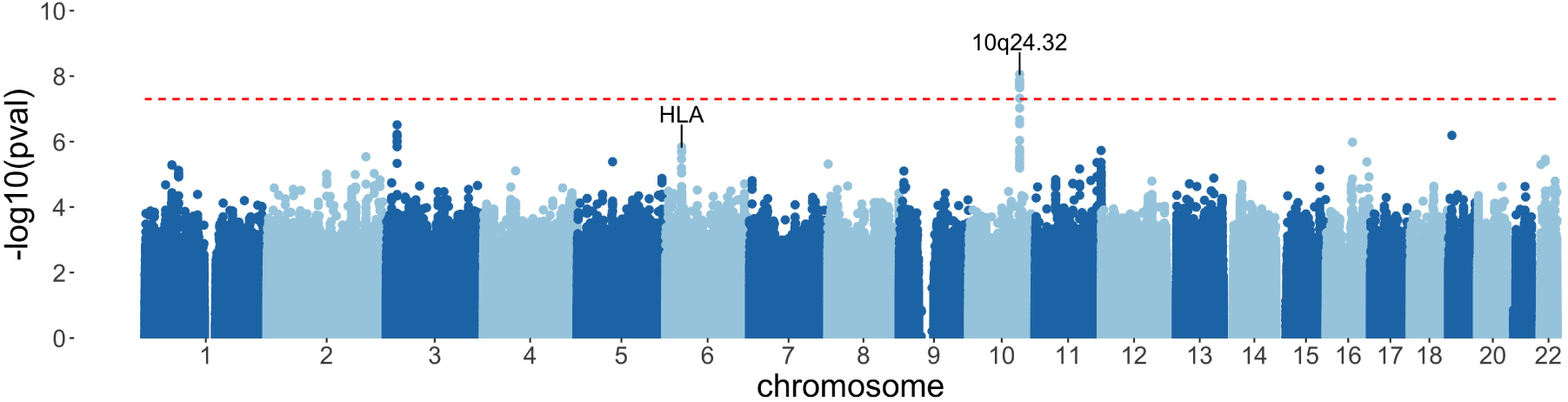
Manhattan plot of leprosy in Malawi and Mali. Evidence for association with leprosy at genotyped and imputed autosomal SNPs and indels (*n* = 9, 616, 523) in Malawi and Mali (492 cases, 639 controls). Association statistics represent a fixed-effects meta-analysis of additive association with disease in Malawi and Mali. The red, dashed line denotes genome-wide significance (*p* = 5 × 10^−8^).

**Figure 2:**
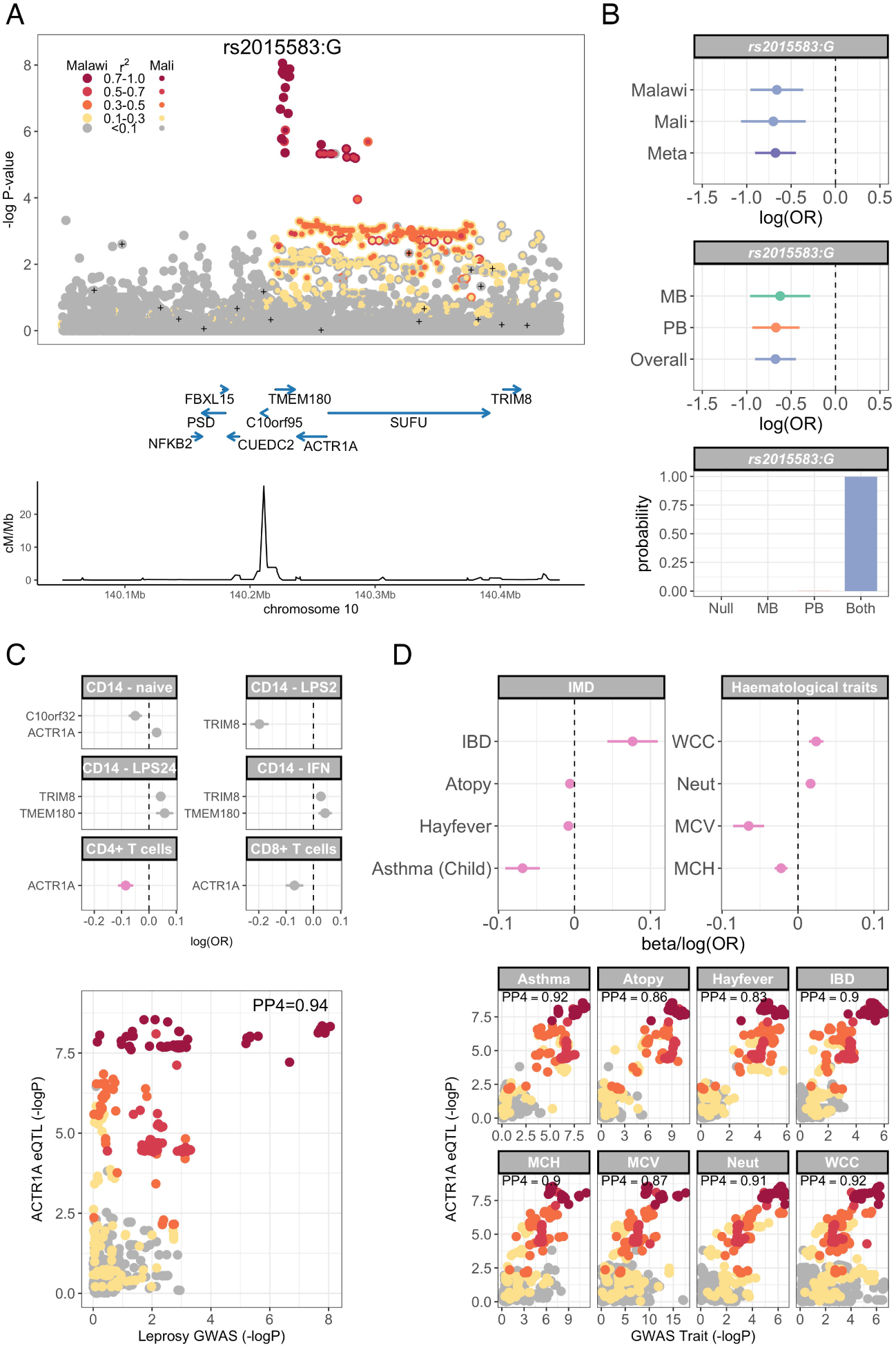
Leprosy association, regulatory function and pleiotropy at chromosome 10q24.32. (A) Regional association plot of leprosy association at chr10q24.32. Association statistics represent a fixed-effects meta-analysis of additive association with disease in Malawi and Mali. SNPs are coloured according to linkage disequilibrium to rs2015583, and genotyped SNPs marked with black plusses. (B) Log-transformed odds ratios and 95% confidence intervals of rs2015583 association with leprosy in Malawi and Mali (top) and stratified by multibacillary and paucibacillary disease (middle). Posterior probabilities of models of rs2015583 association with leprosy: “Null”, no association with leprosy; “MB”, non-zero effect in multibacillary leprosy alone; “PB”, non-zero effect in paucibacillary leprosy alone; “Both”, the same non-zero effect is shared by individuals with multibacillary and paucibacillary leprosy. (C) Log-transformed odds ratios and 95% confidence intervals of rs2015583 association with gene expression in primary immune cells (top). Associations which colocalize with the leprosy association signal (*ACTR1A* expression in CD4^+^ T cells) are highlighted in pink. The *ACTR1A* eQTL in CD4+ T cells colocalizes with the risk locus for leprosy at chr10q24.32 (bottom). SNPs are coloured according to linkage disequilibrium to rs2015583 as above. (D) Log-transformed odds ratios and 95% confidence intervals of rs2015583 association (top) with immune-mediated diseases (IBD, inflammatory bowel disease; atopy; hayfever; childhood-onset asthma) and hematological indices (WCC, white cell count; Neut, neutrophil count; MCV, mean corpuscular volume; MCH, mean corpuscular hemoglobin). The *ACTR1A* eQTL in CD4^+^ T cells colocalizes with the GWAS locus for each trait at chr10q24.32 (bottom). SNPs are coloured according to linkage disequilibrium to rs2015583 as above.

Fine-mapping of the leprosy association at chr10q24.32 identifies a credible set of 19 SNPs with a 95% probability of containing the causal variant, spanning a 6kb region: chr10:104,226,830-104,232,809 (Supplementary Table 2). Genetic variation at this locus has not been previously described as a determinant of leprosy risk. In keeping with the observation that the human genetics of leprosy risk is characterized by considerable heterogeneity of effect between populations, there is no evidence for leprosy association (*p* < 0.05) in Chinese populations (8,156 cases, 15,610 controls) at rs2015583 (*p* = 0.587, *OR* = 0.98) or at any of the 19 variants in the credible SNP set (Dr Z Wang and Prof. F Zhang, personal communication, November 2019).

### A leprosy risk locus modifies *ACTR1A* expression in CD4^+^ T cells

Trait-associated genetic variation identified by GWAS are highly enriched for regulatory variation. To explore whether leprosy-associated genetic variation at chr10q24.32 modifies leprosy risk through its effect on gene expression, we investigated whether the leprosy risk locus colocalizes with expression quantitative trait loci (eQTL) in a range of primary immune cells; monocytes (Fairfax et al., 2014), B cells (Fairfax et al., 2012), NK cells (Gilchrist et al., 2021), neutrophils (Naranbhai et al., 2015), CD4^+^ T cells and CD8+ T cells (Kasela et al., 2017). These data demonstrate colocalization of the leprosy risk locus at chr10q24.32 with an eQTL for *ACTR1A* expression in CD4^+^ T cells (posterior probability of colocalization, *PP* 4 = 0.94), but not gene expression in other cell types (Fig. 2C, Supplementary Table 3). The leprosy protective allele, rs2015583:G, is associated with reduced *ACTR1A* expression in CD4+ T cells (*p* = 4.69 × 10^−9^, *β* = *-*0.085).

### *ACTR1A* expression has pleiotropic effects on risk of immune-mediated disease

A key of feature of genetic determinants of leprosy described to date has been the identification of pleiotropy at leprosy risk loci with other immune-mediated diseases, in particular inflammatory bowel disease (Jostins et al., 2012; Sun et al., 2016). To explore whether leprosy-associated genetic variation modifying *ACTR1A* expression is a determinant of other immune-mediated diseases in human populations, we assessed evidence of colocalization at the CD4^+^ T cell *ACTR1A* eQTL with immune-mediated diseases (n=29) and haematological traits (n=26). In that analysis (Fig. 2D, Supplementary Table 4), in addition to leprosy, the *ACTR1A* eQTL in CD4^+^ T cells colocalizes with four immune-mediated diseases (inflammatory bowel disease, atopy, childhood-onset asthma and hayfever), and four haematological traits (white cell count, neutrophil count, mean corpuscular volume and mean corpuscular haemoglobin). Decreased *ACTR1A* expression in CD4^+^ T cells is associated with perturbations of haematological indices (increased neutrophil and white cell counts, decreased mean corpuscular haemoglobin and volume), increased risk of inflammatory bowel disease and decreased risk of atopy (atopy as a composite trait, childhood-onset asthma and hayfever).

### Replication of leprosy HLA associations

The observation that class I and II HLA alleles are key determinants of leprosy risk has been highly reproducible across diverse populations (Gzara et al., 2020; Wang et al., 2016; Wong et al., 2010).

Motivated by this, and by evidence of association in the HLA observed in our data (Fig. 1), we explored evidence for leprosy association in the HLA in Malawi and Mali at the level of SNPs and classical HLA alleles. In a fixed-effects meta-analysis of leprosy association in Malawi and Mali (Fig. 3A, Supplementary Table 5), the peak classical allele association is a class II allele: HLA-DQB1*04:02 (*p* = 6.74 × 10^−5^, *FDR* = 0.0063, *OR* = 2.1 95% CI 1.75 *-* 2.51). We also observed a leprosy association in the class I region, at HLA-B*49:01 (*p* × 6.02 × 10^−4^, *FDR* = 0.0156), which is independent of HLA-DQB1*04:02 (Supplementary Fig. 3). No significant residual associations were observed after conditioning on both DQB1*04:02 and HLA-B*49:01 (Supplementary Fig. 4). There is no evidence for heterogeneity of effect between populations at HLA-DQB1*04:02 or HLA-B*49:01 (heterogeneity *p* = 0.8836 and *p* = 0.7049, Fig. 3B), and the data best supports a model in which both HLA-DQB1*04:02 and HLA-B*49:01 modify risk of both paucibacillary and multibacillary leprosy (log10 Bayes factors = 2.22 and 0.43 respectively, Fig. 3C,D).

**Figure 3:**
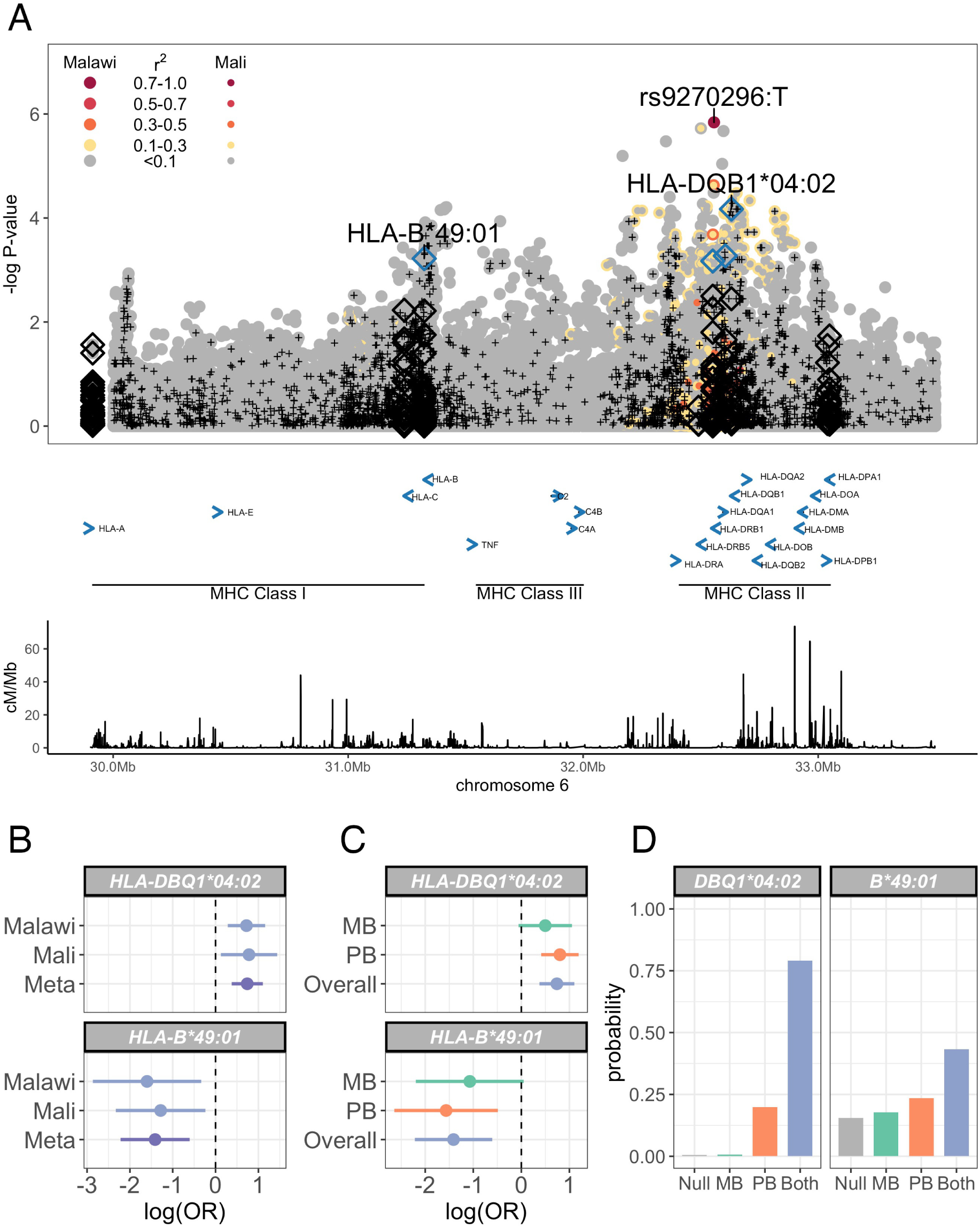
MHC leprosy association in Malawi and Mali. (A) Regional association plot of leprosy association across the HLA region. Association statistics represent a fixed-effects meta-analysis of additive association with disease in Malawi and Mali. SNPs are coloured according to linkage disequilibrium to rs9270926, and genotyped SNPs marked with black plusses. Imputed classical HLA alleles are plotted as diamonds, with significantly associated (FDR < 0.05) alleles highlighted in blue. (B) Log-transformed odds ratios and 95% confidence intervals of HLA-DBQ1*04:02 and HLA-B*49:01 associations with leprosy in Malawi and Mali. (C) Log-transformed odds ratios and 95% confidence intervals of HLA-DBQ1*04:02 and HLA-B*49:01 associations with leprosy stratified by multibacillary and paucibacillary disease. (D) Posterior probabilities of models of HLA-DBQ1*04:02 and HLA-B*49:01 associations with leprosy: “Null”, no association with leprosy; “MB”, non-zero effect in multibacillary leprosy alone; “PB”, non-zero effect in paucibacillary leprosy alone; “Both”, the same non-zero effect is shared by individuals with multibacillary and paucibacillary leprosy.

### Replication of known leprosy associations outside the HLA

Our identification of a genetic variant modifying leprosy risk in African populations, but not in Chinese populations, highlights the inter-population heterogeneity that has been observed across large-scale genetic studies of leprosy susceptibility (Gzara et al., 2020; Wang et al., 2016; Wong et al., 2010). Our study has adequate power (*>* 80%) to replicate (*p* < 0.05) findings at 7 of 34 previously-published leprosy risk loci outside the HLA (Supplementary Table 6). We were able to replicate leprosy associations at 2 loci (Fig. 4A); a missense SNP in *LACC1*, rs3764147 (*p* = 0.004, *OR* = 1.36 95% CI 1.10 *-* 1.67), and a missense SNP in *SLC29A3*, rs780668 (*p* = 0.034, *OR* = 1.28 95% CI 1.02 *-* 1.60). There is no evidence for heterogeneity of effect between populations at rs3764147 or rs780668 (heterogeneity *p* = 0.444 and *p* = 0.159, Fig. 4A), and the data best supports a model in which both rs3764147 and rs780668 modify risk of both paucibacillary and multibacillary leprosy (log10 Bayes factors = 1.64 and 0.68 respectively, Fig. 4B,C). Among the 6 loci at which we were not able to demonstrate replication of previously-published leprosy association despite adequate study power, there is no evidence that our lack of replication reflects effects restricted to multibacillary or paucibacillary disease (Supplementary Table 7). We further considered whether our failure to replicate previously-reported leprosy associations could represent differential linkage disequilibrium to an undefined causal locus between study populations. To test this, we examined evidence for leprosy association within 250kb of each previously-reported leprosy risk locus outside the HLA. In that analysis we identified a promoter variant in *RAB32*, rs34271799, with suggestive evidence of association with leprosy risk in Malawi and Mali (*p*_*MALAW I*_ = 0.0023, *p*_*MALI*_ = 0.0034, *p*_*COMBINED*_ = 6.00 × 10^−5^; *OR* = 0.42, 95%*CI* = 0.28 *-* 0.64, Supplementary Fig. 5). There was no evidence suggestive of leprosy association (*p* < 1 × 10^−4^) within 250kb of any other previously identified leprosy risk locus.

**Figure 4:**
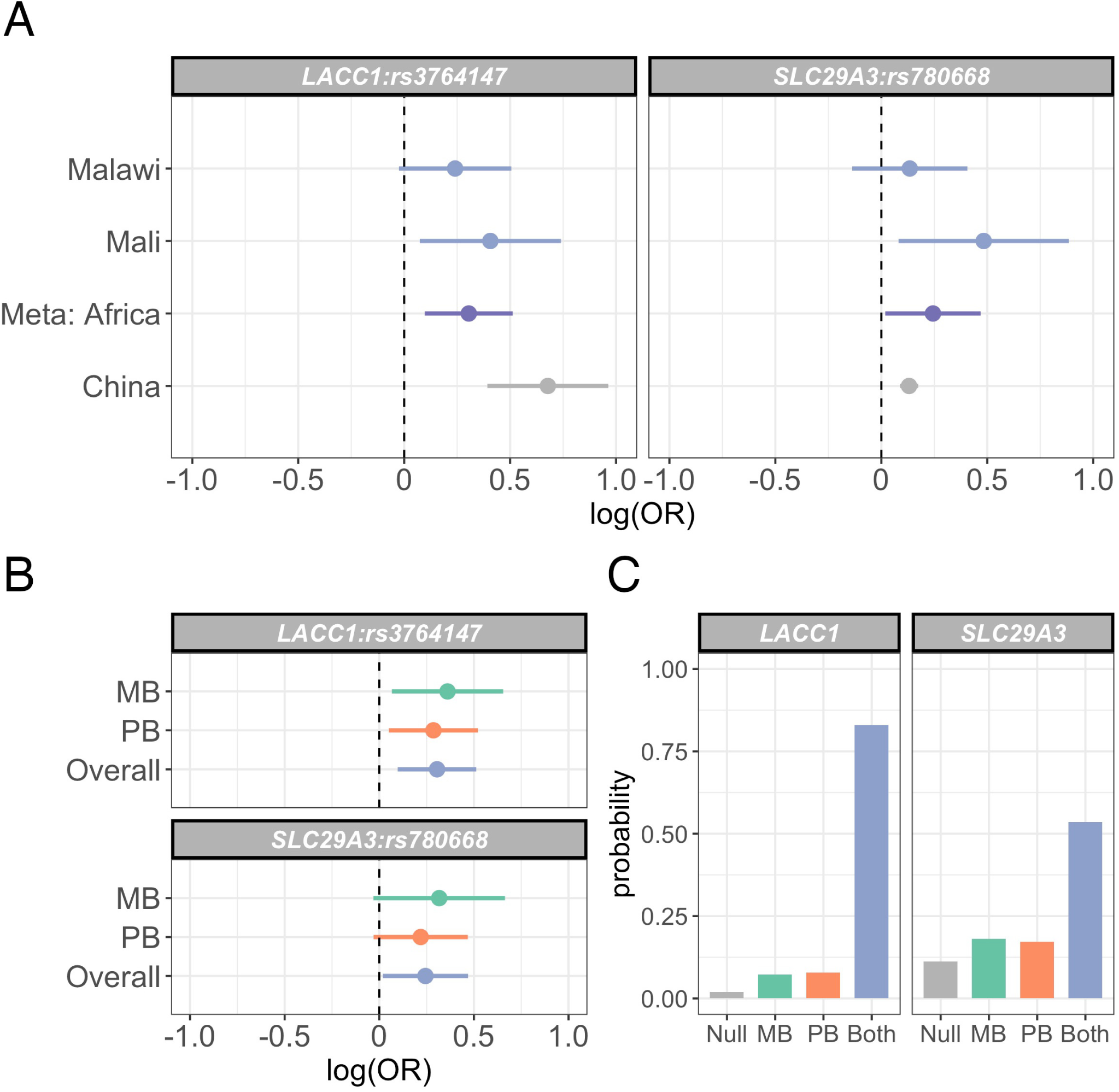
Replication of leprosy associations at LACC1 and SLC29A3 in Malawi and Mali. (A) Log-transformed odds ratios and 95% confidence intervals of rs3764147 and rs780668 associations with leprosy in Malawi and Mali. (B) Log-transformed odds ratios and 95% confidence intervals of rs3764147 and rs780668 associations with leprosy stratified by multibacillary and paucibacillary disease. (C) Posterior probabilities of models of rs3764147 and rs780668 associations with leprosy: “Null”, no association with leprosy; “MB”, non-zero effect in multibacillary leprosy alone; “PB”, non-zero effect in paucibacillary leprosy alone; “Both”, the same non-zero effect is shared by individuals with multibacillary and paucibacillary leprosy.

## Discussion

In this study, we demonstrate that genetic variation at chromosome 10q24.32 is a determinant of leprosy risk in African populations. In common with many examples of trait-associated genetic variation identified by GWAS (Maurano et al., 2012), variation at 10q24.32 modifies risk of leprosy through regulatory effects on gene expression, specifically *ACTR1A* expression in CD4^+^ T cells. We expand upon this, identifying evidence of pleiotropy at 10q24.32, demonstrating a shared genetic architecture of leprosy, inflammatory bowel disease and atopy at this locus. Furthermore, we replicate previously identified leprosy susceptibility loci at *LACC1, SLC29A3*, and with HLA Class I and II alleles.

*ACTR1A* encodes actin-related protein-1 (ARP1), a component of the dynactin complex. Dynactin interacts with the cytoplasmic motor, dynein, facilitating intracellular trafficking of a wide range of intracellular cargos (Roberts et al., 2013; Urnavicius et al., 2015). In T cells specifically, dynactin/dynein complexes direct the accumulation of TCRs and secretory vesicles at the immunological synapse (Hashimoto-Tane et al., 2011; Nath et al., 2016), and are required for nuclear translocation of NF*κ*B in response to T cell stimulation (Shrum et al., 2009). The identification of a CD4^+^ T cell-specific eQTL as a determinant of leprosy reflects the established role of T cell-mediated immunity in leprosy biology. The spectrum of leprosy disease is defined by host T cell responses, with tuberculoid disease being characterised by robust CD4^+^ T cell IFN*γ* responses and patients with lepromatous disease failing to mount anti-*M. leprae* cell-mediated responses (Yamamura et al., 1992). Similarly, leprosy reversal reactions, characterised by painful inflammation of leprosy lesions, are association with infiltration of IFN*γ* producing CD4^+^ T cells (Britton, 1998). In addition, the highly reproducible association between MHC class II alleles and leprosy strongly suggests a role for *M. leprae* antigen presentation as a determinant of leprosy susceptibility *per se*. Our data are complementary to these observations, suggesting a model in which CD4^+^ T cell activation, determined at the level of the T cell as well as that of the antigen presenting cell, modifies susceptibility to leprosy.

The identification of a CD4^+^ T cell-specific eQTL as a risk locus for inflammatory bowel disease and atopy is consistent with the known biology of autoimmune and atopic disease. GWAS-identified risk loci for both autoimmune and atopic disease are highly enriched for regulatory variation active in CD4^+^ T cells, in particular regulatory CD4^+^ T cells (Bossini-Castillo et al., 2019). Our data provide further support for the observation that leprosy and inflammatory bowel disease have a shared genetic architecture (Jostins et al., 2012). We expand on this observation, suggesting a model in which selection pressure imposed by *M. leprae* has shaped the evolution of atopic disease and autoimmune disease in modern populations.

Our identification of a genetic locus modifying leprosy susceptibility in African populations, but with no effect on leprosy risk in well-powered GWAS in Chinese populations, is consistent with the existence of heterogeneity of genetic architecture of leprosy risk between populations. In keeping with this, we observe no evidence of leprosy association in Malawi or Mali at 6 of the 7 non-HLA loci at which we have adequate study power to assess this. We hypothesized that some of these inter-population differences may reflect differential effects of genetic risk loci on multibacillary and paucibacillary disease, however we find no evidence to support this in our data. Differential linkage disequilibrium between assayed variation and a shared causal locus may explain some of the observed inter-population genetic heterogeneity of leprosy risk. In keeping with this we observe modest evidence of leprosy association at *RAB32*, which is distinct from that reported in Chinese populations. Understanding whether genetic variation at *RAB32* is associated with leprosy risk in African populations, and whether this is distinct from that observed in Chinese populations, will require replication in additional study populations.

Here we define regulatory variation at *ACTR1A* as a novel determinant of leprosy susceptibility in African populations. Moreover, regulatory variation at *ACTR1A* has pleiotropic effects on hematological indices in European populations and risk of IBD and atopy. A shared genetic architecture for leprosy and IBD has been previously described. We expand on this, strengthening the evidence that selection pressure driven by leprosy has shaped the evolution of immune-mediated disease in modern populations. Our colocalization analyses identify *ACTR1A* as a potential therapeutic target for autoimmune and atopic disease, and deepens our understanding of leprosy biology, which will be key in informing the development of novel control strategies. More broadly, our data highlights the importance of defining the genetic architecture of disease across genetically diverse populations, and that disease insights derived from GWAS in one population may not readily translate to all affected populations.

## Materials and methods

### Ethics and consent

All cases and controls were recruited following informed consent of the participant or their parent/guardian. The study protocol detailing recruitment and sample collection within KPS, Malawi was approved by the National Health Sciences Research Committee of Malawi and by the Ethics Committee of the London School of Hygiene and Tropical Medicine. The study protocols detailing recruitment and sample collection at the Institut Marchoux, Mali and at the Centre Hospitalier Universitaire Gabriel Toure, Bamako, Mali were approved by the University of Bamako Ethics Review Board. Genotyping and imputation for additional Malian controls were collected as part of the MalariaGEN project, for which the study protocol was reviewed by Oxford University Tropical Research Ethics committee (OXTREC), Oxford, UK (OXTREC 020–006). The overall study design, including re-appraisal of study samples using genome-wide genotyping, was reviewed and approved by Oxford University Tropical Research Ethics committee (OXTREC), Oxford, UK (OXTREC 560-15).

### Study Participants

Leprosy case and control samples were recruited to the study in Karonga, Malawi and Bamako, Mali. Participant recruitment in Malawi (Fitness et al., 2004; Wallace et al., 2004) and Mali (Meisner et al., 2001) have been described previously. In brief, cases of leprosy were diagnosed in both settings on the basis of clinical examination, split skin smear and histopathologic examination of biopsy material. Adults or children with “certain” or “probable” cases were considered eligible for recruitment (Ponnighaus et al., 1987). Cases were further defined as having paucibacillary (PB) or multibacillary (MB) disease on the basis of clinical examination and bacteriological index *>* 1 on slit-skin smear or biopsy.

In Malawi, cases (n=327) and controls (n=436) were recruited to the study within the KPS, a longterm community-based, epidemiological study in Northern Malawi (Ponninghaus et al., 1987). Leprosy cases were identified through active population surveys in the 1980s, followed by enhanced passive case detection in the 1990s. Control samples were individuals within the KPS with no history or clinical features of leprosy, matched to case samples with respect to age, sex and geographic area of residence.

In Mali, between April and June 1997, patients with leprosy (n=247) presenting to Institut Marchoux, Bamako, Mali (formerly Mali’s national leprology center now Hôpital Dermatologique de Bamako) were recruited to the study as described previously (Meisner et al., 2001). Control participants (n=185), following exclusion of leprosy by clinical examination and history, were recruited in the same study setting among hospital staff and patients with diagnoses other than leprosy. In addition, we supplemented control numbers in the Malian replication study using healthy control samples (n=183), also recruited in Bamako, collected as part of the MalariaGEN project (Malaria Genomic Epidemiology, 2019; Toure et al., 2012).

Cases in Malawi were recruited at a median age of 47 years (range 15 to 83 years) and 187 were female (57%). Among leprosy cases in Malawi, 47 had multibacillary disease and 275 paucibacillary disease. Control samples in Malawi were recruited at a median age of 44 years (range 15 to 82 years) and 273 were female (63%). Cases in Mali were recruited at a median age of 45 years (range 13 to 85 years) and 136 (55%) were female. Among leprosy cases in Mali, 165 had multibacillary disease and 82 paucibacillary disease. Control samples in Mali were recruited at a median age of 30 years (range 14 to 72 years) and 113 were female (62%). Additional MalariaGEN control samples were recruited at a median age of 3 years (range 0 to 15 years) and 91 (50%) were female.

### Genotyping

Genomic DNA was extracted from study samples as described previously (Fitness et al., 2004; Meisner et al., 2001). Following quantification (Ahn et al., 1996), samples were genotyped using the Illumina African Diaspora Power Chip platform (Johnston et al., 2017) and genotypes called using GenCall in GenomeStudio (Illumina). Using consensus strand information from the array manifest file and a remapping pipeline (Dr William Rayner, Wellcome Centre for Human Genetics, Oxford) we aligned genotypes such that all alleles are on the forward strand. Throughout genetic positions reflect GRCh37.

### Sample quality control

We calculated per sample quality control (QC) metrics in PLINK (Purcell et al., 2007). For each sample we calculated the proportion of missing genotype calls, heterozygosity and the mean X and Y channel intensities. We plotted mean X and Y channel intensities (Supplementary Fig. 6) and missingness against heterozygosity (Supplementary Fig. 7), defining outlier samples using ABERRANT (Bellenguez et al., 2012). We used PLINK to estimate sample sex from genotype data, excluding samples with discordant genotype and metadata sex information. To identify duplicated and related samples, we calculated pairwise relatedness between samples in PLINK. We considered samples with relatedness *>* 0.75 to be duplicates, and additionally identified samples with relatedness *>* 0.2. In both cases we retained the sample with the highest genotyping call rate of a duplicated/related sample pair. We calculated principal components (PC) in EIGENSTRAT (Price et al., 2006). To identify population outliers, we plotted study sample PCs against a background of African Genome Variation Project (Gurdasani et al., 2015) samples, identifying outliers by visual inspection (Supplementary Fig. 8). For both PC and relatedness computations we used an LD-pruned SNP set with regions of high linkage disequilibrium (LD) excluded. The first two major principal components differentiate self-reported ethnicity in both Malawi (Supplementary Fig. 9) and Mali (Supplementary Fig. 10).

### SNP quality control

Prior to genome-wide imputation, we extracted genotypes from non-duplicated, autosomal SNPs and applied the following SNP QC filters; SNP missingness *>* 10%, minor allele frequency (MAF) < 1%, Hardy-Weinberg equilibrium (HWE) *p* < 1 × 10^−20^ and plate effect *p* < 1 × 10^−6^. HWE was calculated among control samples for each cohort. Plate effect represents an association test of nondifferential missingness with the plate on which each sample was genotyped.

### Imputation

Following sample (Supplementary Table 8) and SNP (Supplementary Table 9) QC, genotypes at 351,236 autosomal SNPs from 612 samples (Malawi) and genotypes at 367,433 autosomal SNPs from 350 samples (Mali) were taken forward for phasing and genome-wide imputation. We performed phasing of genotypes in SHAPEIT2 (Delaneau et al., 2012), phasing genotypes across each chromosome for all samples from each country jointly. We then imputed untyped autosomal genotypes using IMPUTE2 (v2.3.0) (B. Howie et al., 2011; B. N. Howie et al., 2009), in 5Mb chunks using the 1000 Genomes Phase III as a reference panel. We used 250kb buffer regions and effective sample size of 20,000.

### HLA imputation

We used HLA*IMP:03 (Motyer et al., 2016) to impute classical HLA alleles into our datasets. As input to HLA*IMP we used genotypes passing sample and SNP QC thresholds in the HLA region (chr6:28Mb-36Mb). HLA*IMP:03 uses a multi-population reference panel, including individuals of African ancestry. HLA imputation performed well, with estimated imputation accuracies ranging between 95% and 99.8%. We took forward HLA alleles present in both Malawi and Malawi (MAF *>* 1%), including 93 classical alleles (42 class I and 51 class II) in downstream analysis.

### Additional cross-platform quality control

We noted that relatively few Mali control samples (n=142) were available for inclusion in the association analysis. To address this, we used genotypes from additional control samples (n=183) of representative ethnicity collected as part of the MalariaGEN project (Malaria Genomic Epidemiology, 2019; Toure et al., 2012). These samples have been genotyped on the Illumina Omni 2.5M platform. Sample genotypes have been phased using SHAPEIT2 (Delaneau et al., 2012) and untyped genotypes imputed genome-wide using IMPUTE2 (v2.3.2) (B. Howie et al., 2011; B. N. Howie et al., 2009) with 1000 Genomes Phase III as a reference panel. The SNP and sample QC used in processing these samples (Malaria Genomic Epidemiology, 2019) is highly analogous to the QC we applied to our study samples. MalariaGEN SNP QC excluded poorly genotyped SNPs using the following metrics; SNP missingness (thresholds 2.5-10% dependent on study population), MAF < 1%, HWE *p* < 1 × 10^−20^, plate effect *p* < 1 × 10^−3^ and a recall test quantifying changes in genotype following a re-clustering process *p* < 1 × 10^−6^. MalariaGEN sample QC excluded samples prior to imputation according to the following metrics; channel intensity, missingness, heterozygosity (outlier thresholds determined by ABERRANT (Bellenguez et al., 2012)), population outliers and duplicated samples (relatedness *>* 0.75). Of note, related samples (relatedness > 0.2) are retained for imputation purposes.

We defined a shared subset of SNPs genotyped and passing SNP QC on both platforms (*n* = 26, 136), from which we calculated relatedness estimates and PCs. We removed one of related pairs (relatedness 0.2) from the MalariaGEN samples, which resulted in a final sample size of 519 (208 cases, 311 controls). Inspection of the PCs demonstrate no further population outliers, and that the 10 major PCs are nondifferential with respect to genotyping array (Supplementary Fig. 10).

### Association analysis

Following imputation, SNPs were taken forward for association analysis if they passed the following QC metrics; MAF *>* 4%, imputation info score *>* 0.4, HWE *p* < 1 × 10^−10^. For the Mali samples, these QC filters were applied overall and for samples genotyped on each genotyping platform individually. At each variant passing QC we tested for association with leprosy case-control status in a logistic regression model in SNPTEST (Marchini et al., 2007) in each cohort. At loci of interest, we used multinomial logistic regression, implemented in SNPTEST, to estimate the effect of the genetic variation on leprosy risk stratified by multibacillary and paucibacillary disease. We used control status as the baseline stratum and cases of multibacillary and paucibacillary leprosy as strata. To account for confounding variation, in particular population structure, we included the six major principal components of genotyping data in all models. In addition, in Mali, we included genotyping platform as an additional categorical covariate. At variants passing QC thresholds in both cohorts, we then performed genome-wide meta-analysis under a frequentist fixed-effects model using BINGWA (Band et al., 2015). For association analysis using HLA allele imputations we coded posterior probabilities of each HLA allele to represent carriage of 0, 1 or 2 copies of that allele. Association analysis and meta-analysis was performed in SNPTEST and BINGWA as above. For HLA association analysis we corrected for the number of classical alleles tested (n=93) and considered FDR < 0.05 to be significant.

### Bayesian comparison of models of association

We compared models of association at loci of interest with multibacillary and paucibacillary leprosy, as estimated by multinomial logistic regression, using a Bayesian approach. We considered four models of effect, defined by the prior distributions on the effect size:

“Null”: effect size = 0, i.e. no association with leprosy.

“MB”: effect size *N* (0, 0.2^2^) for multibacillary disease, but no effect in paucibacillary disease.

“PB”: effect size *N* (0, 0.2^2^) for paucibacillary disease, but no effect in multibacillary disease.

“Both”: effect size *N* (0, 0.2^2^) and fixed between multibacillary and paucibacillary disease (*ρ* = 1).

For each model we calculated approximate Bayes factors (Wakefield, 2009) and posterior probabilities, assuming each model to be equally likely a priori. Statistical analysis was performed in R.

### Definition of credible SNP sets

We used a Bayesian approach to identify a set of SNPs with 95% probability of containing the causal locus at the leprosy susceptibility locus at chr10q24.32. Approximate Bayes’ factors (Wakefield, 2009) were calculated for each SNP in the region (a 200kb surrounding rs2015583) with a prior distribution of *N* (0, 0.2^2^). All SNPs were considered equally likely to be the causal variant a priori. A set of SNPs with 95% probability of containing the causal SNP was defined as the smallest number of SNPs for which the summed posterior probabilities exceed 0.95.

### eQTL mapping and colocalization analysis

We used the R package coloc (Giambartolomei et al., 2014) to identify evidence of causal variants shared by eQTL in primary immune cells and GWAS-identified trait associated loci (including leprosy). Coloc adopts a Bayesian approach to compare evidence for independent or shared association signals for two traits at a given genetic locus. We tested for colocalization between leprosy susceptibility at the chr10q24.32 locus and previously-published eQTL mapping studies in naïve and stimulated primary immune cells from individuals of European ancestry (Fairfax et al., 2014; Fairfax et al., 2012; Gilchrist et al., 2021; Kasela et al., 2017; Naranbhai et al., 2015); NK cells (n = 245), B cells (n = 283), monocytes (n = 414), CD4^+^ T cells (n = 293), CD8^+^ T cells (n = 283), neutrophils (n = 101), LPS-stimulated monocytes (2 hours, n = 261; 24 hours, n = 322) and IFN*γ*-stimulated monocytes (n = 267). We considered evidence for colocalization for each gene within a 250kb window of the peak leprosy association (rs2015583). We considered a posterior probability *>* 0.8 supporting a shared causal locus to be significant.

To assess evidence for pleiotropy with other disease traits we again used coloc to test for the presence of a shared causal locus between the *ACTR1A* eQTL in CD4^+^ T cells and 55 GWAS traits (Supplementary Table 4); hematological indices (n = 26) and immune-mediated diseases (n = 13) from the UK Biobank (http://www.nealelab.is/uk-biobank/, accessed 26th March 2021), immune-mediated diseases from the NHGRI-EBI GWAS Catalog (n = 13, ftp://ftp.ebi.ac.uk/pub/databases/gwas/summary statistics/, accessed 26th March 2021), and inflammatory bowel disease traits from the International Inflammatory Bowel Disease Genetics Consortium (n = 3, https://www.ibdgenetics.org/downloads.html, accessed 26th March 2021).

## Data Availability

On publication, genotype and phenotype data for Malawian and Malian cases and controls will be made available via the European Genotype-Phenome Archive. Genotype and phenotype data describing the MalariaGEN Malian samples have been deposited in the European Genome-Phenome Archive (EGA; study accession EGAS00001001311). Access to MalariaGEN datasets on EGA is by application to an independent data access committee. On publication, a full set of association summary statistics will be made available for download through the NHGRI-EBI GWAS Catalog (https://www.ebi.ac.uk/gwas/downloads/summary-statistics).

## Data Availability

On publication, genotype and phenotype data for Malawian and Malian cases and controls will be made available via the European Genotype-Phenome Archive. Genotype and phenotype data describing the MalariaGEN Malian samples have been deposited in the European Genome-Phenome Archive (EGA; study accession EGAS00001001311). Access to MalariaGEN datasets on EGA is by application to an independent data access committee. On publication, a full set of association summary statistics will be made available for download through the the NHGRI-EBI GWAS Catalog (https://www.ebi.ac.uk/gwas/downloads/summary-statistics).

## Acknowledgements

This publication uses genotyping data from the MalariaGEN consortial project, as described in Malaria Genomic Epidemiology Network, et al. Nature Communications, 2019 (https://doi.org/10.1038/s41467-019-13480-z). We thank Stuart Mucklow and Giles Warner for their assistance in collecting samples in Mali. JJG and AJM are funded by National Institute for Health Research (NIHR) Clinical Lectureships. During this work AVSH was supported by a Wellcome Trust Senior Investigator Award (HCUZZ0) and by a European Research Council advanced grant (294557). The research was supported by the Wellcome Trust Core Award Grant Number 203141/Z/16/Z with additional support from the NIHR Oxford BRC. The views expressed are those of the author(s) and not necessarily those of the NHS, the NIHR or the Department of Health and Social Care.

## Supplementary Figures

**Figure S1:**
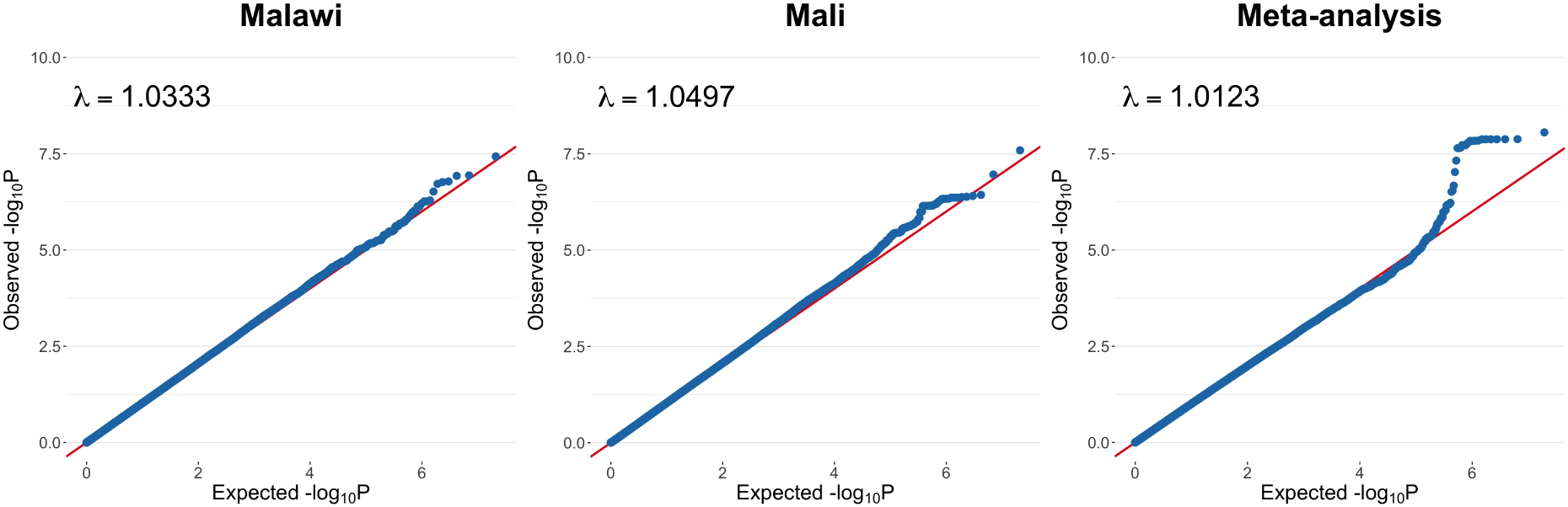
Quantile-quantile plots of leprosy association. QQ-plots of leprosy association in Malawi (284 cases, 328 controls, SNPs = 10,511,695), Mali (208 cases, 311 controls, SNPs = 10,514,676) and fixed-effects meta-analysis of both populations (cases = 492, controls = 639, SNPs = 9,616,523).

**Figure S2:**
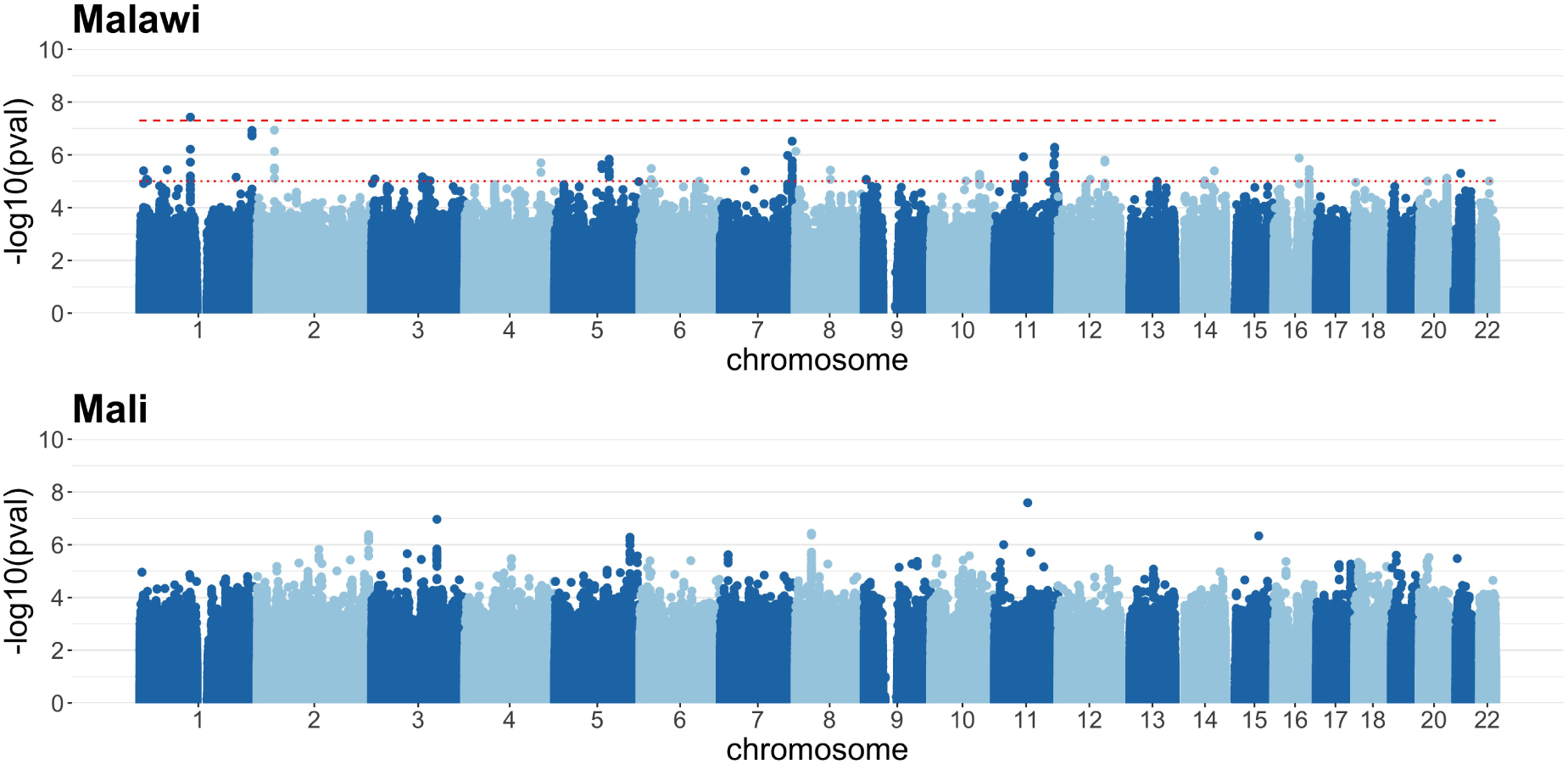
Manhattan plots of leprosy association. Manhattan plots of leprosy association in discovery (Malawi, 284 cases, 328 controls, SNPs = 10,511,695) and replication (Mali, 208 cases, 311 controls, SNPs = 10,514,676) samples. P-value thresholds are annotated on the Malawi Manhattan plot: dashed line, *p* = 1 × 10^−8^ (genome-wide significance); dotted line, *p* = 1 × 10^−5^ (threshold for suggestive association).

**Figure S3:**
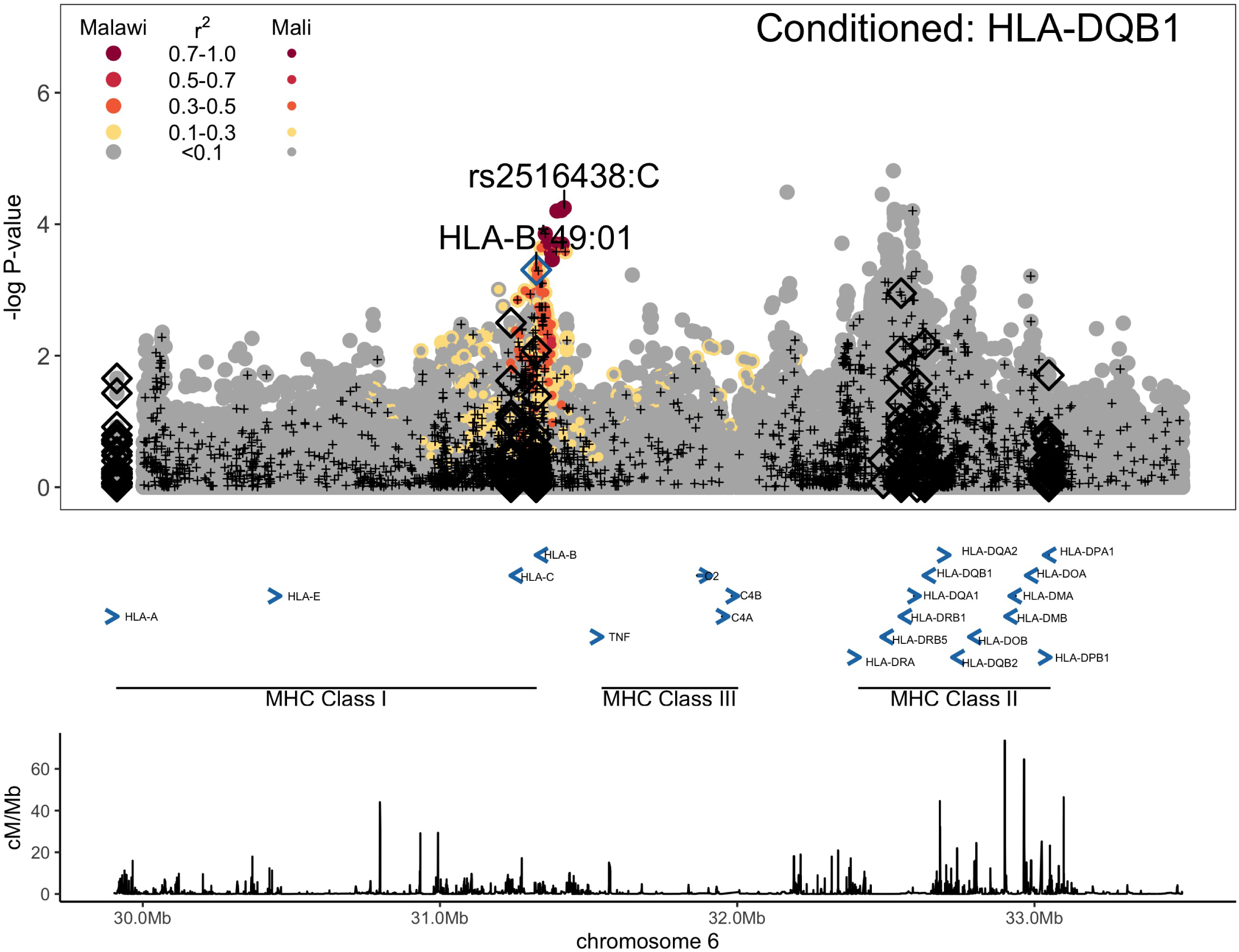
HLA Leprosy association conditioned on HLA-DQB1*04:02. Association statistics represent a fixed-effects meta-analysis of additive association with disease in Malawi and Mali conditioned on HLA-DQB1*04:02. SNPs are coloured according to linkage disequilibrium to rs2516438, and genotyped SNPs marked with black plusses. Imputed classical HLA alleles are plotted as diamonds, with significantly associated (FDR < 0.05) alleles highlighted in blue.

**Figure S4:**
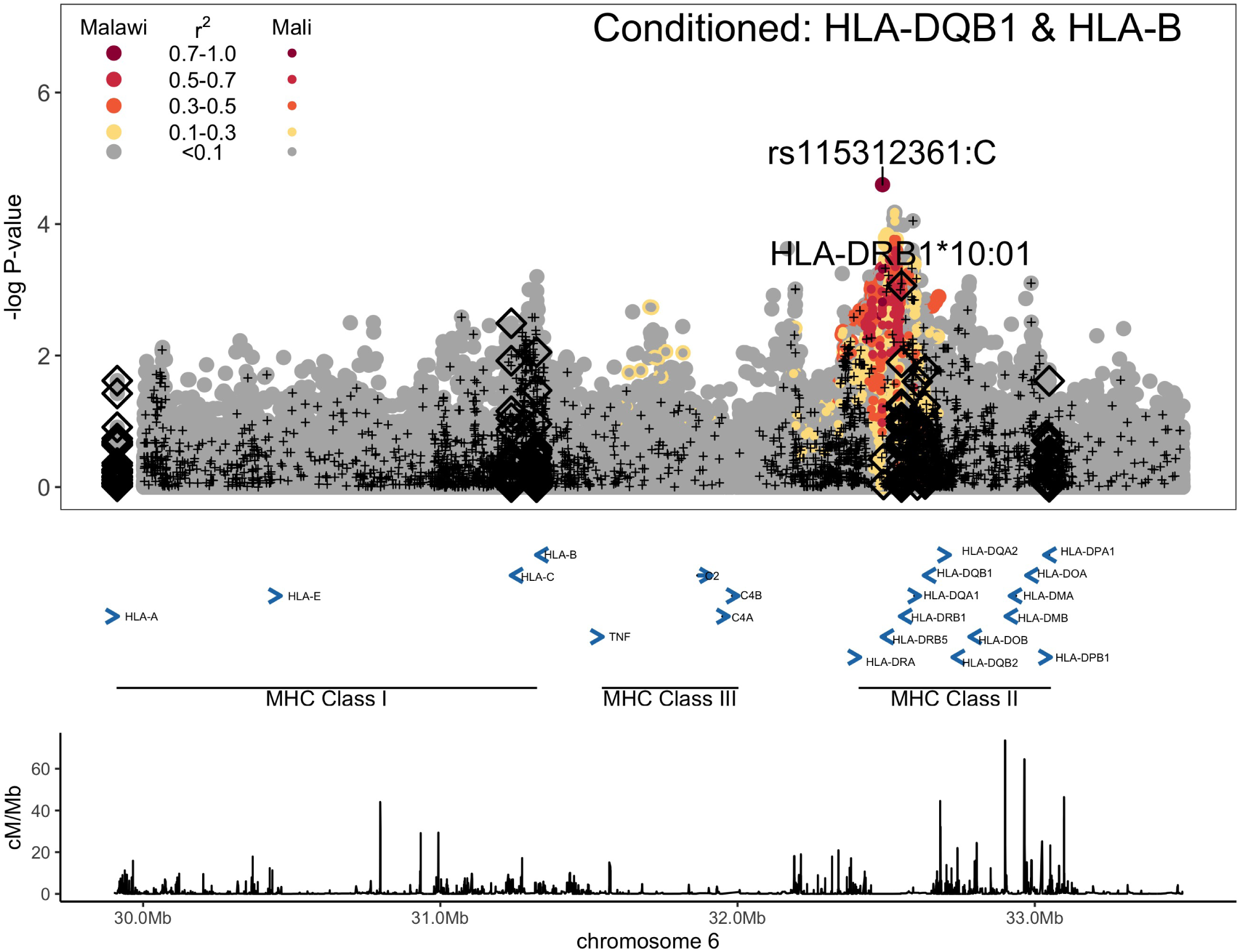
HLA Leprosy association conditioned on HLA-DQB1*04:02 and HLA-B*49:01. Association statistics represent a fixed-effects meta-analysis of additive association with disease in Malawi and Mali conditioned on HLA-DQB1*04:02 and HLA-B*49:01. SNPs are coloured according to linkage disequilibrium to rs115312361, and genotyped SNPs marked with black plusses. Imputed classical HLA alleles are plotted as diamonds. No significantly associated (FDR < 0.05) alleles remain after conditioning on HLA-DQB1*04:02 and HLA-B*49:01.

**Figure S5:**
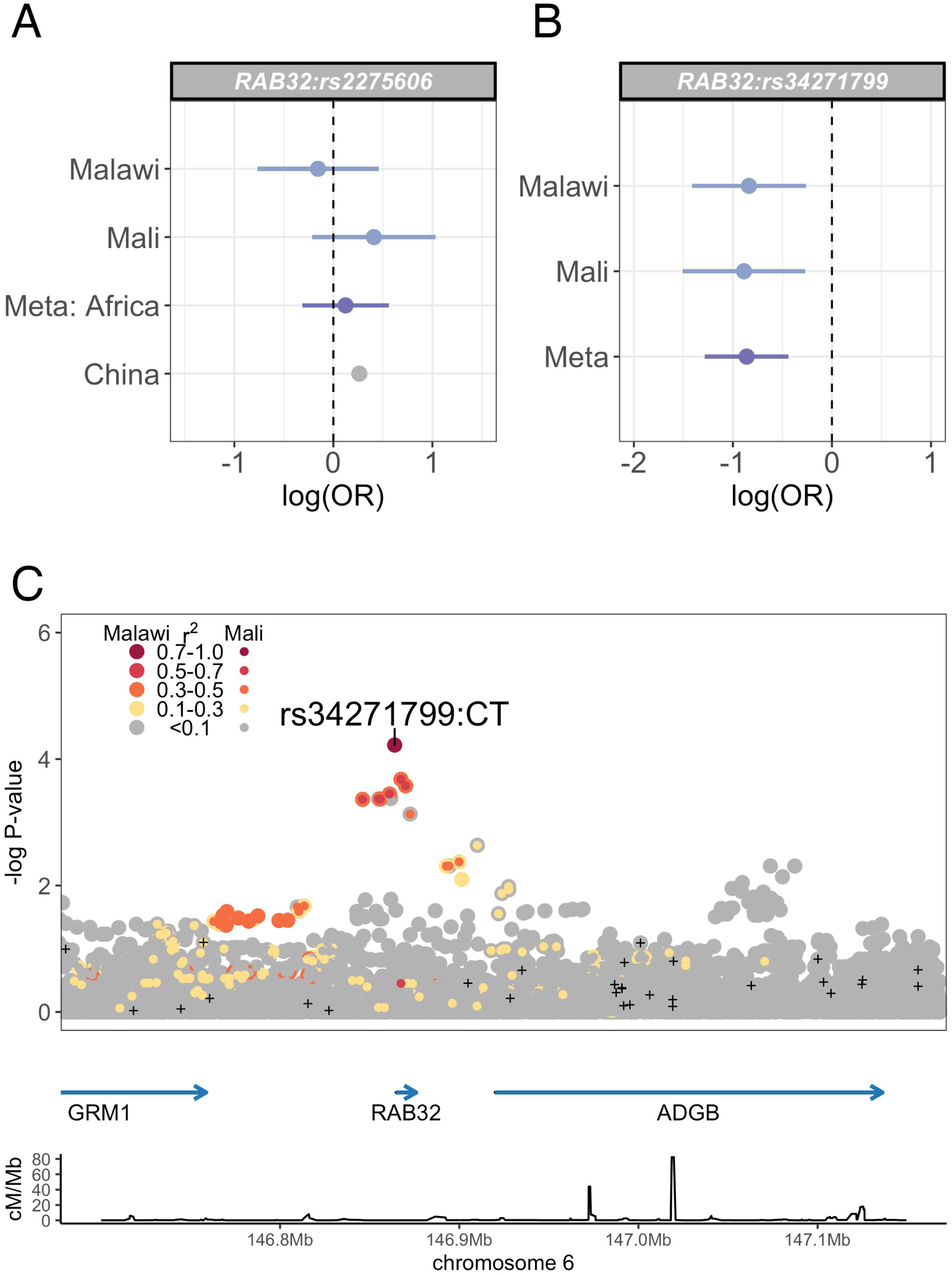
Evidence for leprosy association at the RAB32 locus. (A) Log-transformed odds ratios and 95% confidence intervals of rs2275606 association (peak association in Chinese GWAS data) with leprosy in Malawi, Mali and China. (B) Log-transformed odds ratios and 95% confidence intervals of rs34271799 association with leprosy in Malawi and Mali. (C) Regional association plot of leprosy association at the RAB32 locus. Association statistics represent a fixed-effects meta-analysis of additive association with disease in Malawi and Mali. SNPs are coloured according to linkage disequilibrium to rs34271799, and genotyped SNPs marked with black plusses.

**Figure S6:**
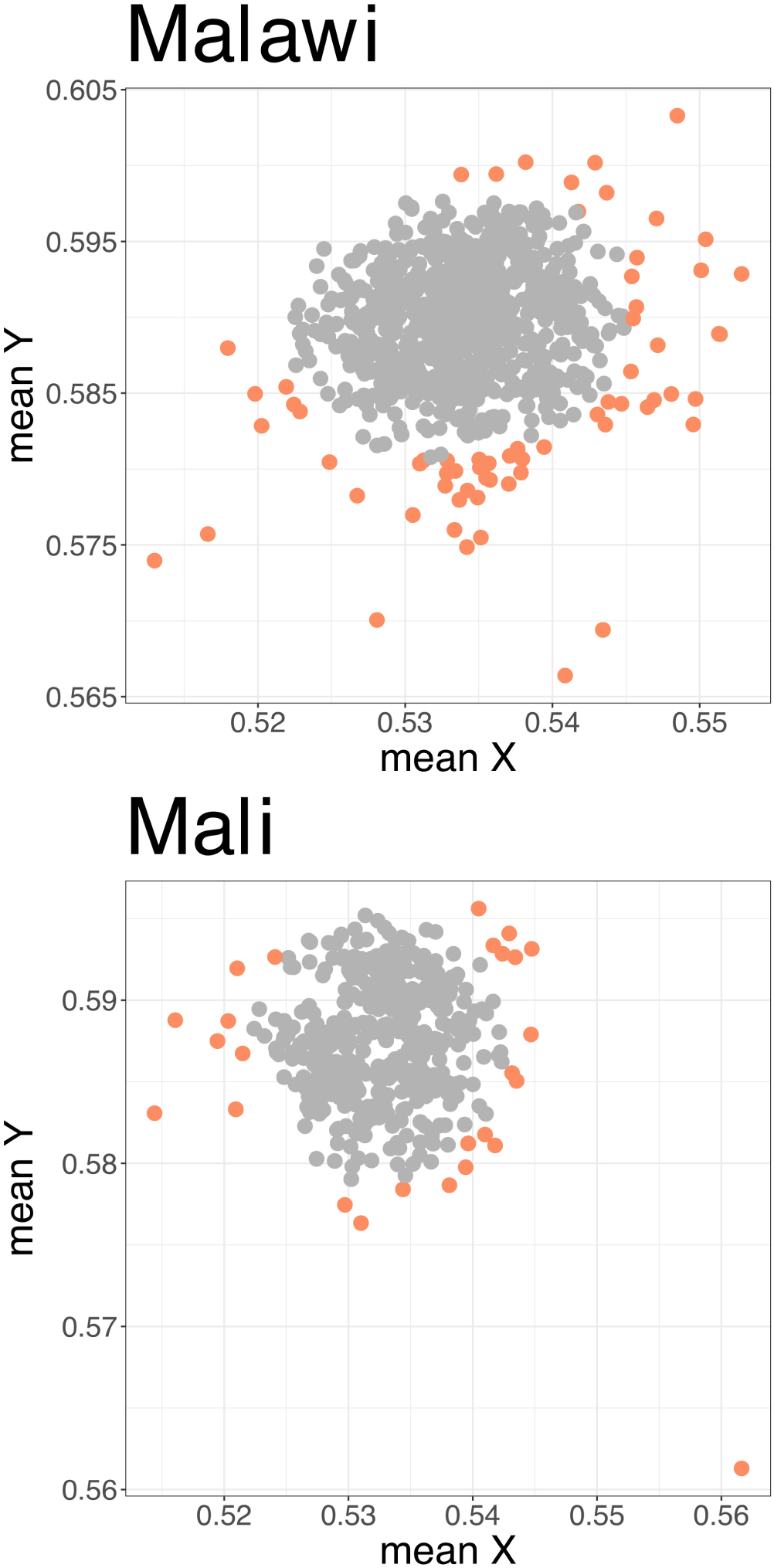
Sample X and Y channel intensities. (A) Mean X and Y channel intensities for Malawi (top) and Mali (bottom) samples. Outlying samples were identified using ABERRANT and are highlighted (orange).

**Figure S7:**
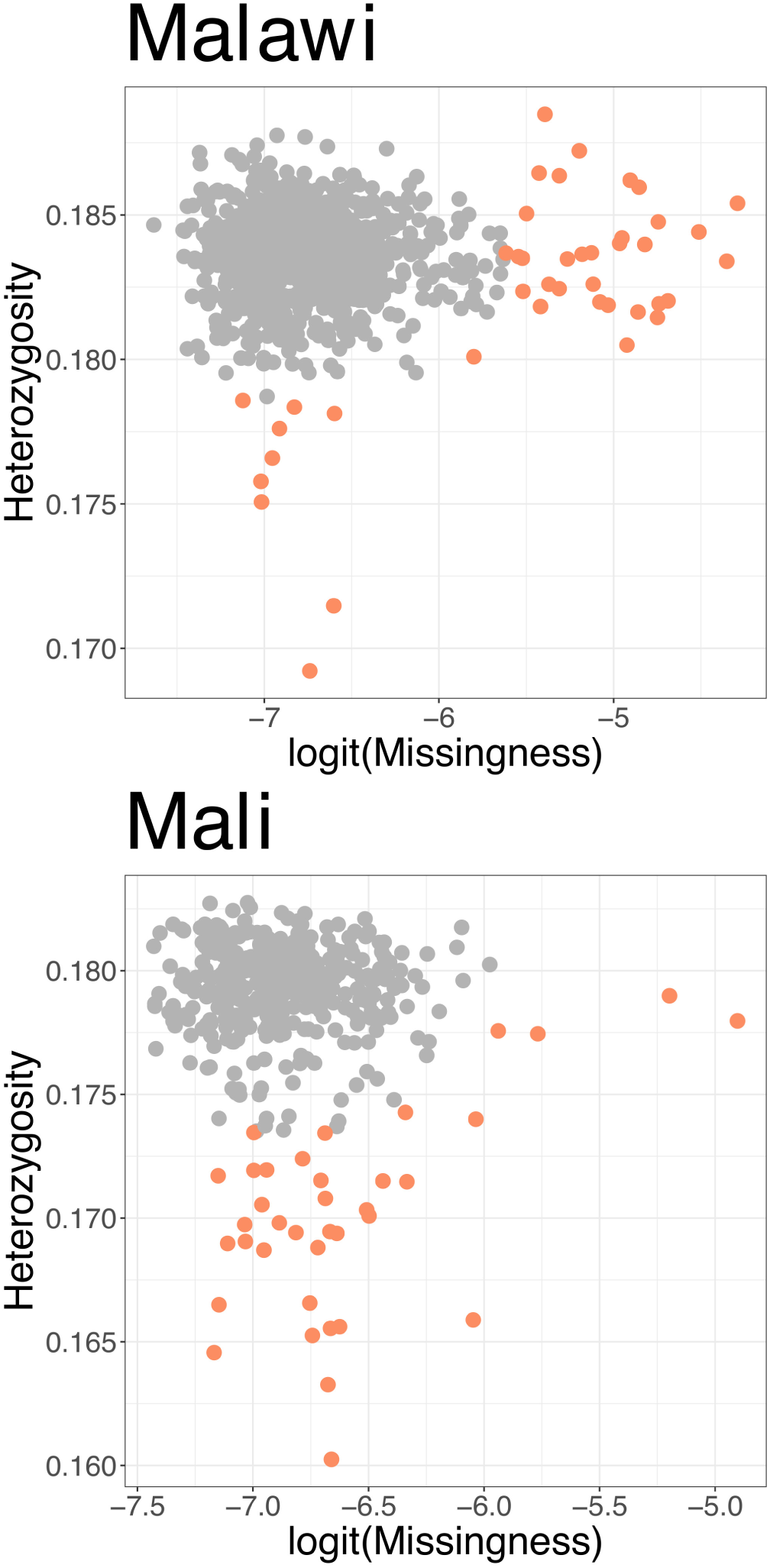
Sample missingness and heterozygosity. (A) Mean sample genotype missingness plotted against heterozygosity for Malawi (top) and Mali (bottom) samples. Outlying samples were identified using ABERRANT and are highlighted (orange).

**Figure S8:**
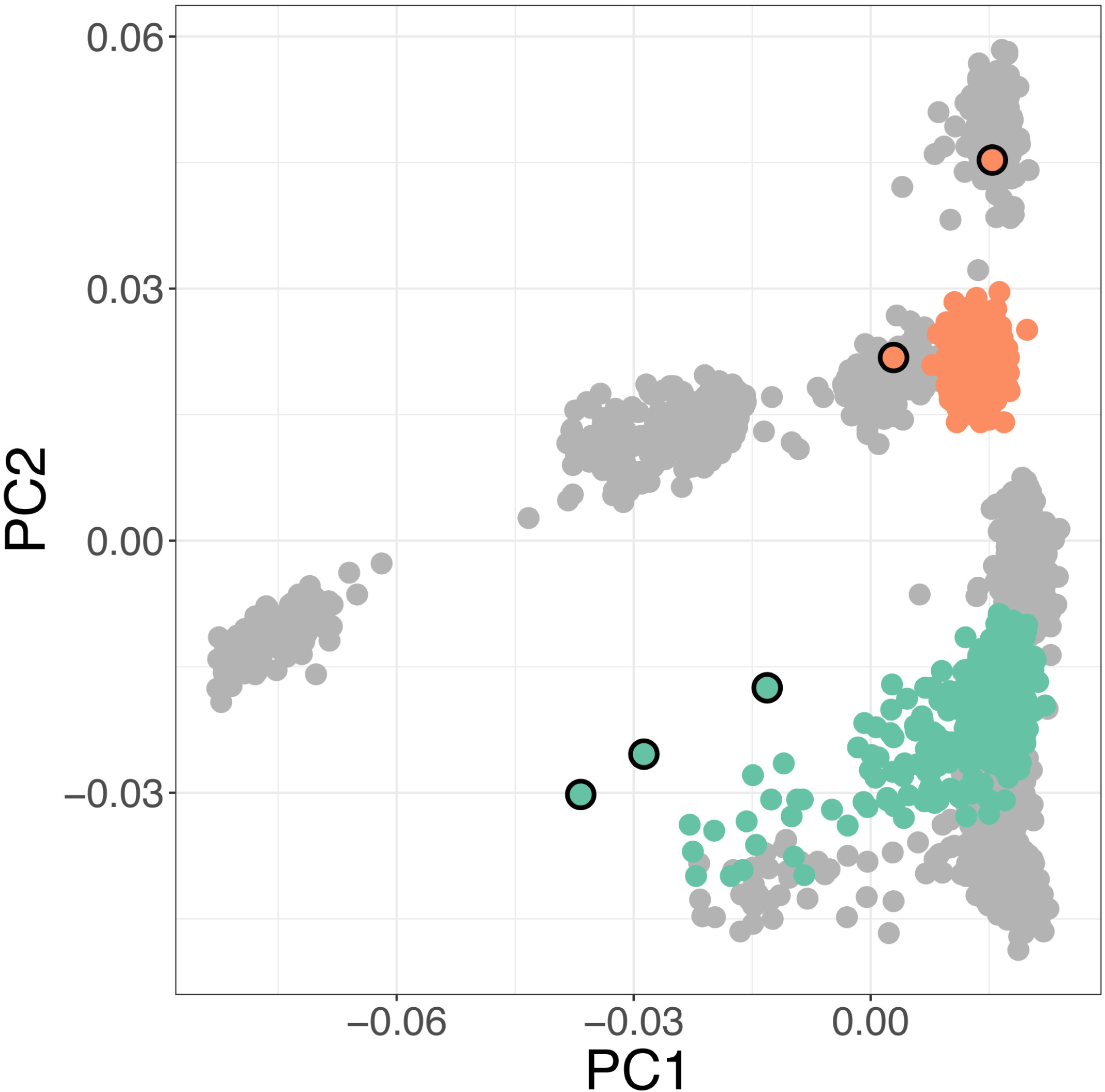
Population outliers. Plot of the major two principal components of genome wide genotyping data. Malawi study samples are plotted in orange and Mali study samples in green, against a background of African Genome Variation Project samples (gray). Outliers are highlighted (black rings).

**Figure S9:**
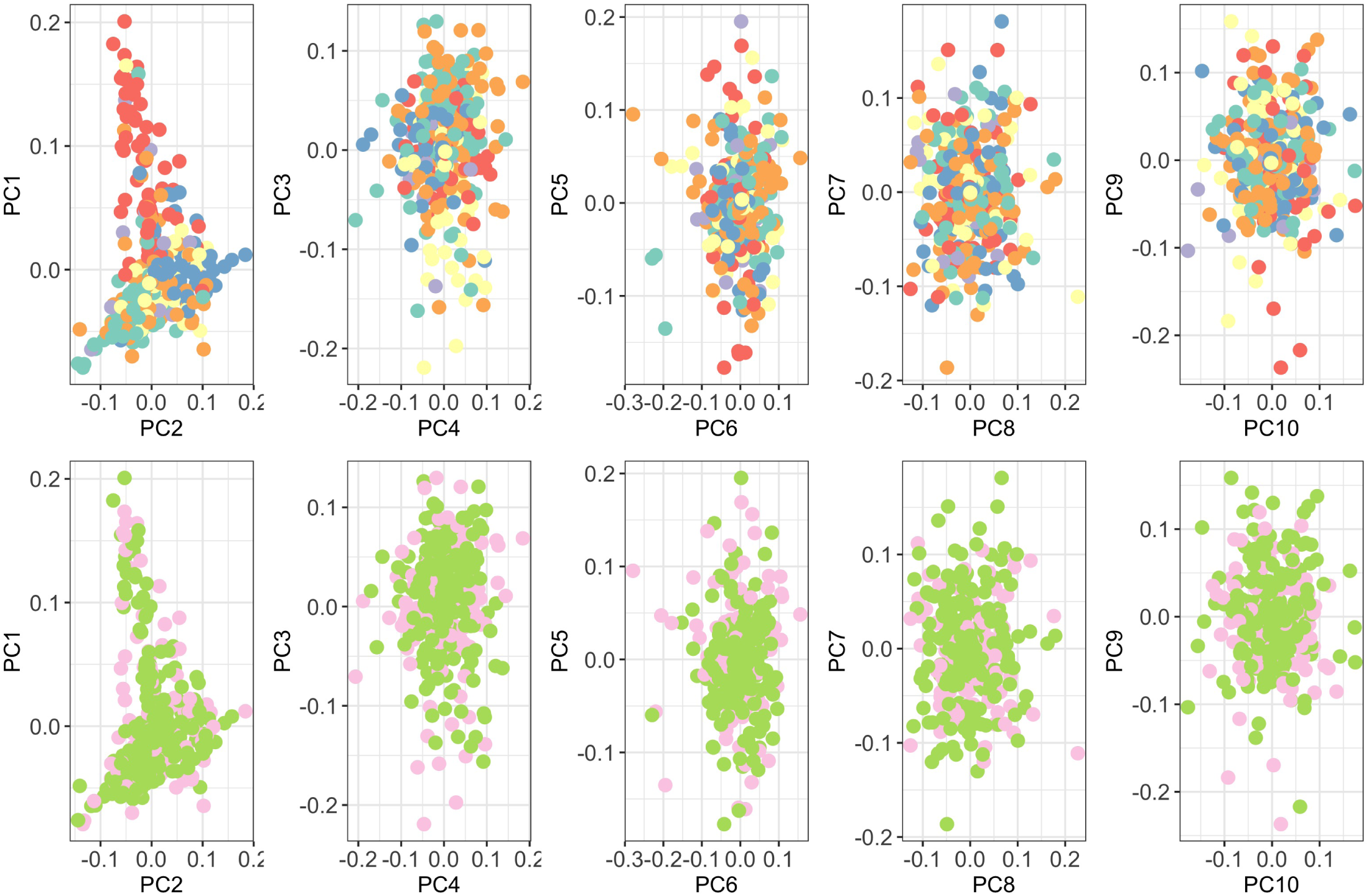
Principal components of Malawian genome-wide genotyping data. Individuals are color-coded according to self-reported ethnicity (top) and case-control status; cases in pink, controls in green (bottom).

**Figure S10:**
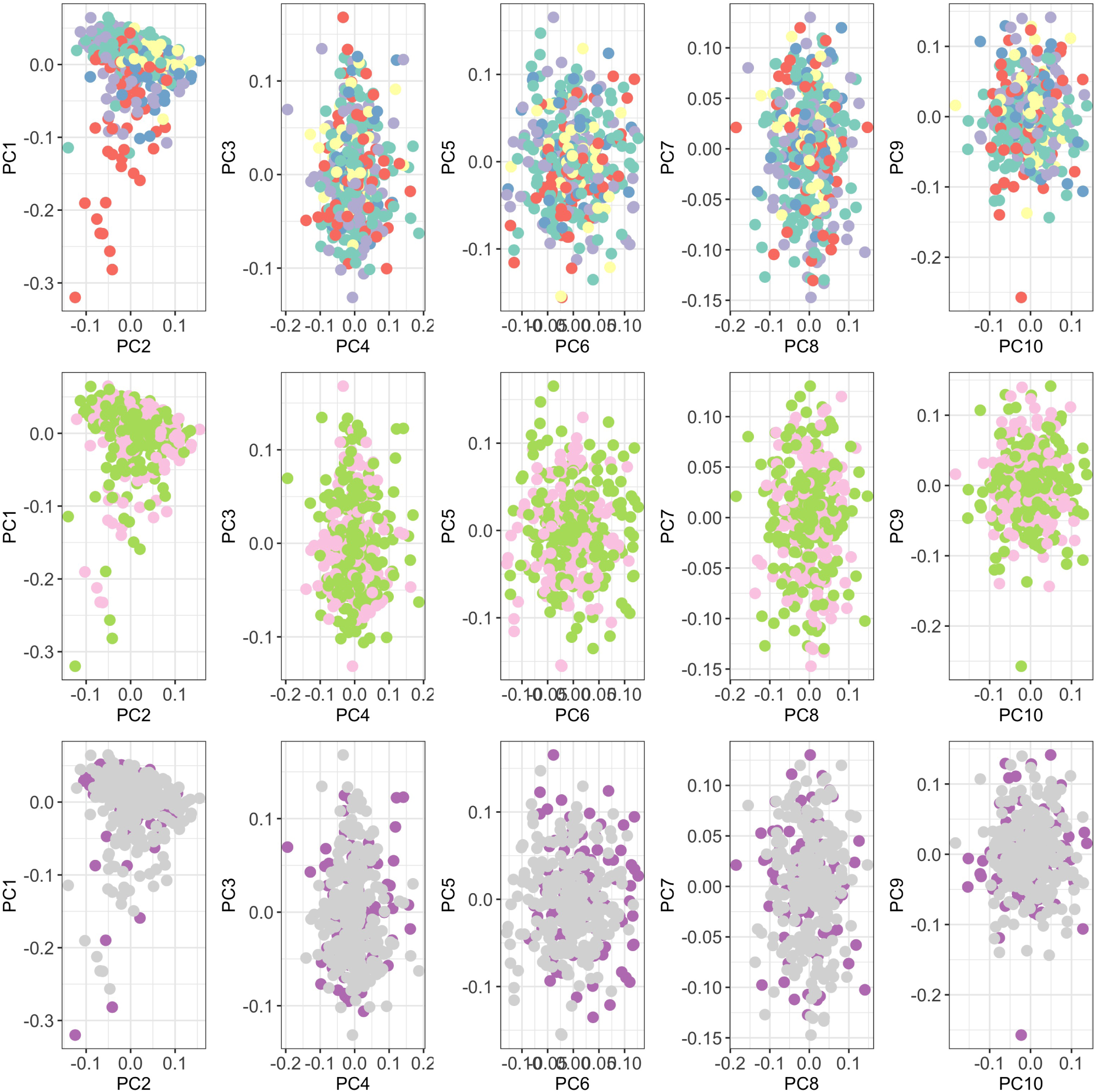
Principal components of Malian genome-wide genotyping data. Individuals are color-coded according to self-reported ethnicity (top), case-control status (middle; cases in pink, controls in green), and genotyping platform (bottom; Omni 2.5M in purple, Africa Diaspora Power Chip in gray).

